# The Build-Up of Aerosols Carrying the SARS-CoV-2 Coronavirus, in Poorly Ventilated, Confined Spaces

**DOI:** 10.1101/2020.08.11.20173195

**Authors:** Björn Birnir

## Abstract

A model of the distribution of respiratory droplets and aerosols by Lagrangian turbulent air-flow is developed and used to show how the SARS-CoV-2 Coronavirus can be dispersed by the breathing of an infected person. It is shown that the concentration of viruses in the exhaled cloud can increase to infectious levels with time (grow linearly), in a confined space where the air re-circulates. The model is used to analyze the air-flow and SARS-CoV-2 Coronavirus build-up in a restaurant in Guangzhou, China [30, 28]. It is concluded that the outbreak of Covid-19 pandemic in the restaurant in January 2020, is due to the build-up of the airborne droplets and aerosols carrying the SARS-CoV-2 Coronavirus and would not have been prevented by standard ventilation. A comparison with standard models for aerosol concentration shows that, in the absence of ventilation, the decay of the aerosol concentration is also controlled by the decay time of the virions in aerosols. This decay time is very long and a steady state is not achieved in the time-frame of the contagion. Instead the concentration exhibits a polynomial increase and reaches infectious levels in a relatively short time, explaining the outbreak in the restaurant in Guangzhou.

## 1 Introduction

We develop a model describing the concentrations of small droplets and aerosols as a function of time. The small droplets quickly evaporate into aerosols and we show that the aerosol concentration may increase linearly in time, with the reinjection of the contaminated air, and not quickly approach a steady state concentration, as predicted by the conventional models.

The understanding of the mechanism of infection by the SARS-CoV-2 Coronavirus has evolved considerably since the beginning of the Covid-19 pandemic. It is believed that the most common form of transmission is by respiratory droplets containing the coronavirus. These droplets were classically divided into heavier droplets that settle at a limited distance around the infected individual, small droplets, 5 − 10 micrometers in diameter, and aerosols (or droplet nuclei) < 5 micrometers in diameter, that can remain airborn for an extended period and are convected by air inside confined spaces. The more recent scientific understanding is more nuanced. The pathogen-containing droplet can be carried by a turbulent cloud, emitted when the infected person coughs or sneezes. The droplets are active particles in this turbulent cloud and interact with the turbulent air and with each other. Experiments show that the cloud can rapidly spread up to a distance of 7-8 meters [12, 39, 14] (up to 25 feet). The droplet cloud, that is carried by air and can be airborne for hours, overlaps the artificial boundary of droplets and aerosols. We will adopt the terminology *droplet/aerosols*, in this paper, for droplets and aerosols around 1 − 10 micrometers in diameter. This term will apply to all droplets and aerosols that are airborne for times greater than 1 − 10 minutes, that carry the coronavirus and are a source of contagion. The small droplets evaporate quickly and turn into aerosols as they are transported by air and interact with the ventilation system, so if we exclude large drops and droplets > 10 micrometers, we will be able to restrict our conclusions to the concentration of aerosols < 5 micrometers in diameter.

There is mounting evidence that droplet/aerosols are an important source of contagion, in confined spaces, see [34, 33, 13, 36, 16, 26]. This makes it important to establish models that allow a computation of the concentration of the droplet/aerosols and how that concentration evolves over time. Such models would allow a computation of the risk of contagion and develop methods for its mitigation. In this paper we develop such a model based on Lagrangian fluid dynamics of turbulent air-flow. Then we use it to analyze an outbreak of infection [30] in a restaurant in Guangzhou, China, and compare with a numerical and experimental study [28] of the same outbreak. We develop a model describing the concentrations of droplet/aerosols as a function of time. These small droplets quickly evaporate into aerosols and we show that the aerosol concentration may increase linearly in time with reinjection of the contaminated air, and not quickly approach a steady state concentration, as predicted by the conventional models. The reason for this is that with poor or non-existent ventilation only the decay of the virions in aerosols controls the decay of concentration. This virion decay time is very long [44]. The Lagrangian theory permits a computation of parameters in this model.

Our study shows that the concentration of the aerosols may increase significantly in a confined space in the span of one hour. This increase in concentration is the likely cause of the outbreak in the restaurant in Guangzhou, China, see [30] and [28]. As discussed in [37] this increased concentration may spread by air-conditioning ducts from one confined space to another. The remedy is to greatly increase the effectiveness of the air-conditioning filters to filter microscopic particles, see [37] and Appendix D, and/or create a cross flow through the confined space that washes the droplet/aerosols away. Applying such methods in confined spaces such as restaurants, offices, buses and classrooms, may dramatically decrease the contagion rates.

## 2 Application to Covid-19

In a recent paper [30] the infection of 3 families by one infected person in an air-conditioned restaurant in Guangzhou, China, is described, see Figure 1. One of the families had travelled from Wuhan and ate lunch in the restaurant where the other two families were present. The first family contained one person (A1) who fell ill later the same day and went to the hospital. 12 days later 9 other members of the three families had fallen ill with Covid-19. The infection is consistent with droplet transmission because no one else at the restaurant nor the servants fell ill. Only the persons in the direct airstream of the air-conditioner fell ill.

**Figure 1:**
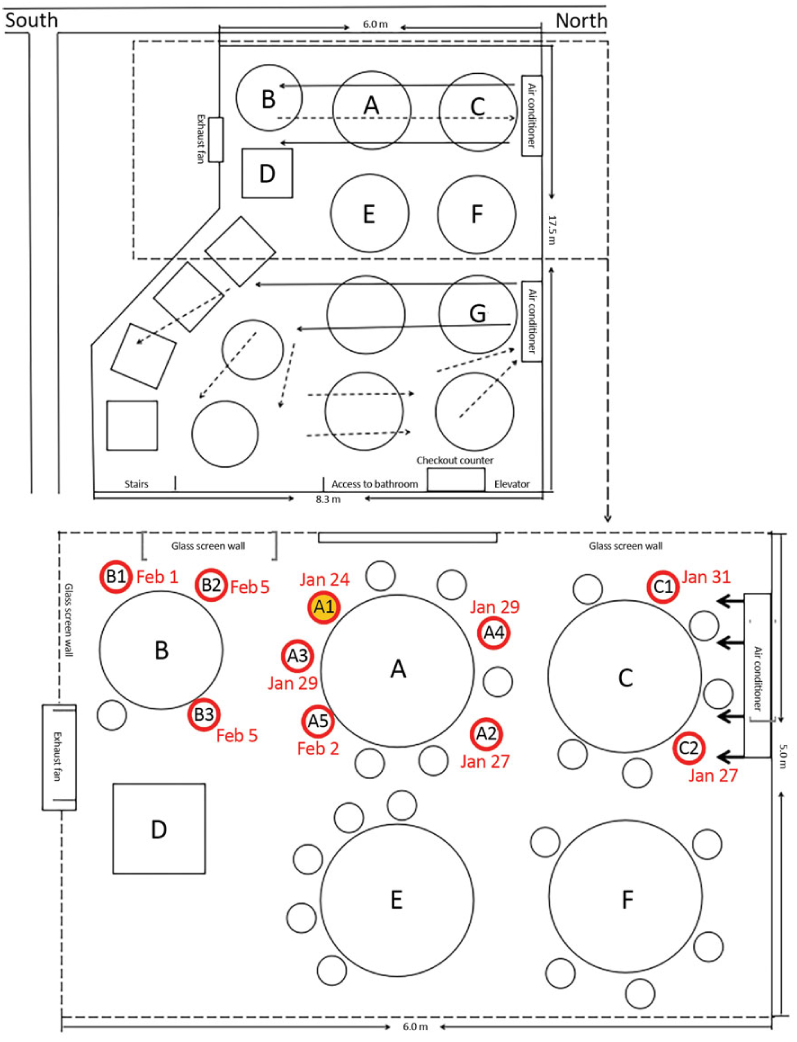
A sketch showing the arrangement of restaurant tables and air conditioning airflow at the site of an outbreak of 2019 coronavirus, in Guangzhou, China, 2020. Red circles indicate where patients that developed the disease were located; yellow-filled red circle (A1) indicates the location of the infected patient. The figure is taken from [30].

In the following analysis we will use the above theory to simulate the dispersion of the droplet/ aerosols in the restaurant. The part of the restaurant where the contagion took place is of dimensions 6 meters length and 3 meters width, see Figure 1. Air-conditioners are set so that the restaurant guest experience wind air blowing at the velocity of 0.25 meters per second, see [1]. The corresponding Taylor-Reynolds number is *Re*_λ_ = 705, since the air is flowing along a 3 meter distance from Table A to the wall where the air-conditioner is located. The three meter distance, perpendicular to the air-flow, is taken to be the distance across the largest table, where the infected person was sitting, and including one half-meter for each person sitting on opposite sites. Using the energy dissipation ε = 1.2, taken from the experimental and simulation data in [6], we get the Kolmogorov time scale τ_η_ = 3.55 ms (milliseconds). We employ the theoretical structure function *S*_2_ in Appendix A and use the stochastic closure theory (SCT), see [7, 8, 10], to compute the coefficient *C*_2_ in the exponential of the formula for the Richardson distribution (B.3). This computation, that interpolates the coefficients computed in [25] for the Reynolds number *Re*_λ_ = 705, is explained in Appendix C.

The results of the simulations for the restaurant in Guangzhou are shown in Figures 2-6. In Figure 2 we show the Richardson distribution as a function of time and space. We assume that the infected person exhales 12 times a minute and each exhale event last 2.4 seconds, or almost half of the breathing period, that is 5 seconds. We are taking twice the lower limit of the breathing rate, that we take to be the shortest time of a breathing event, or the most forceful one (a cough), see [19]. Each time unit is τ_η_ = 3.55 ms, and we are computing 2000 units for a total of 7.1 seconds. In the left figure in Figure 2, we show the complete distribution for one exhaling event, roughly 340 points ≈ 1.2 second. Observe that the distance between the points is greater in the beginning (Richardson dispersion) but becomes smaller after the second half, 1.2 to 2.4 seconds, of the exhale event (Eulerian diffusion). We make this clearer in Figure 7, where on the left we plot the distribution of the cloud for first 340 points or 1.2 seconds and on the right for the subsequent 340 point or the subsequent 1.2 seconds, to see what happens to the initial cloud immediately afterwards. We call the left the Lagrangian part and the right the Eulerian part of the exhale and propagation events, because the first involves Lagrangian and the second Eulerian eddies. The cloud is clearly more diffuse during the Lagrangian part (Richardson dispersion) and more concentrated during the Eulerian part. The concentration of the droplet/aerosols is slightly (1.16 times) greater during the Eulerian phase. The cloud is traveling with the airflow, at 0.25 m/s, so the total length of the cloud is 0.6 meters, the Lagrangian part extends to 0.3 meters and the Eulerian part covers the subsequent 0.3 meters.

**Figure 2:**
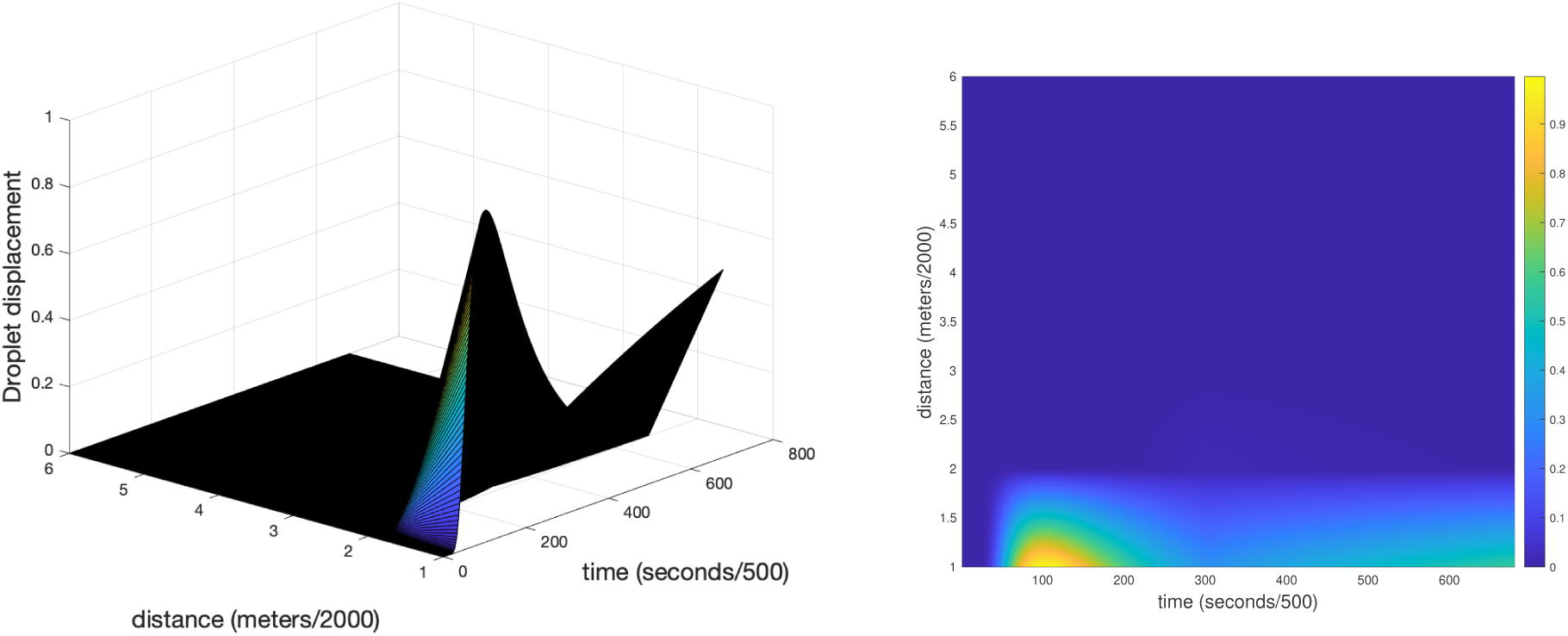
The Richardson distribution for the particle separation.

The Richardson distribution for the particle separation for passive scalars, in Appendix C, allows us to compute the shape and the volume of the cloud of droplet/aerosols exhaled by an infectious person. We assume that the person is stationary (seated) and that he talks to the people around him and breathes in various directions in an 180° angle with him in the center. The velocity of is exhaled breath is taken to be 1.5 meters per second and this is what creates the Lagrangian cloud. This velocity is not sustained because the infected person both exhales and inhales. The cloud extends in the radial (*y*) direction but it is carried by the ambient air flow, that is driven by the poorly ventilated air conditioning, in a fixed direction, at the velocity of 0.25 meters per second. Thus no matter in which direction the puffs of droplet/aerosols were exhaled, they get shaped into a cloud with a cylindrical symmetry by the ventilation wind along the axis (*x*), see Figure 5, and the exhalation velocity along the radial (*y*) direction. Using the Richardson distribution we can compute the shape of the cloud in the radial (*y*) direction and using the velocity of the ambient air flow, in the *x* direction, we compute the extent of the cloud in the *x* direction. The infectious person is sitting with his head in the center of the middle of the cylinder in Figure 5. This geometry allows us to compute the emission rate of the droplet/aerosols below.

It is important to understand that the Richardson distribution is not giving the instantaneous shape of an exhaled cloud. The exact shape of the exhaled cloud will vary from one exhale event to another, a reflection of the fact that these are stochastic processes. But a large number of these events are taking place, 12 per minute, and the Richardson distribution is producing the shape of the averaged cloud, averaged over all of these exhale events. Moreover, a person does not get infected by small droplet and aerosols from a single exhale event. Infection results from an exposure to a large number of exhale events over time, see [22]. Thus the Richardson distribution is exactly giving us the information that we need.

One can make a rough estimate of how the concentration of the droplet/aerosols builds up in time using the above theory and the CFD simulations shown in Figure 4. The volume of the whole space containing the contamination is 3 by 6 by 3.14 meters = 56.52 *m*^3^, see [28]. A pyramid with base on the wall opposite the air-conditioner mostly contains the blue contamination cloud, see Figure 4. Thus the cloud does not fill the available space but only this pyramid with the volume 56.52*/*3 = 18.84 *m*^3^. The radius of a cylinder containing the exhaled cloud, is the height of the cloud, and reaches its maximum at 100 units on the left figure in Figure 7. This corresponds to 0.35 seconds and the velocity determining the spread is probably not the velocity at which the cloud is drifting but the velocity at which the droplets-aerosols are being ejected. This is taken to be 1.5 m/s in [28] and we will use the same value here. (Recall that there are two air velocities involved: the velocity of the exhaled air, in the radial *y* direction, and the ambient wind velocity, in the *x* direction, see Figure 5.) This give the maximum radius 1.5 times 0.35 = 0.53 meters. Computing the area under the curves (blue and red) in Figure 3 (left), and rotating it around the *x* axis, we get the volume, 0.132 m^3^. The cloud has a vase shape, see Figure 3 (left), but its volume is equivalent to the volume of a cylinder of radius 0.264 meters and length 0.6 meters, see Figure 5. If we let the exhaled clouds be contained in a cylinder with this radius extending the length of the contamination area we get the volume 1.32 *m*^3^ for this cylinder. Namely, the length of the cylinder, 6 meters, is 10 times the length of the exhaled cloud, 0.6 meters. The droplet/aerosols in the cylinder now get spread to the pyramid, by the air-conditioning and heat conduction, see Figure 5. This decreases their concentration by a factor of 18.48*/*1.32 = 14. However, the infected person keeps exhaling a new droplet/aerosol cloud every 5 seconds and since the extent of these clouds is 0.6 meters he needs to exhale 10 clouds to fill the 6 meter long cylinder. This takes 50 seconds or 0.83 minutes. In one hour he has filled the cylinder 72 = 60/0.83 times. Thus with no droplet/aerosols lost, the exhaust fan was closed see [28], the resulting concentration in the pyramid is 72 × 1*/*14 = 5.16 times what it was in the cylinder, the first time it was full. In other words, the concentration for everyone sitting at the three tables A, B and C, in Figure 1, in less than 12 minutes, is what it would be if they were sitting next to the infected person. In one hour, the concentration is more than five times what it was initially, if every person on the three table was sitting next to the infected person. This build-up of the concentration of the droplet/aerosols containing the SARS-CoV-2 Coronavirus is the likely reason for the outbreak in the restaurant in Guangzhou.

**Figure 3:**
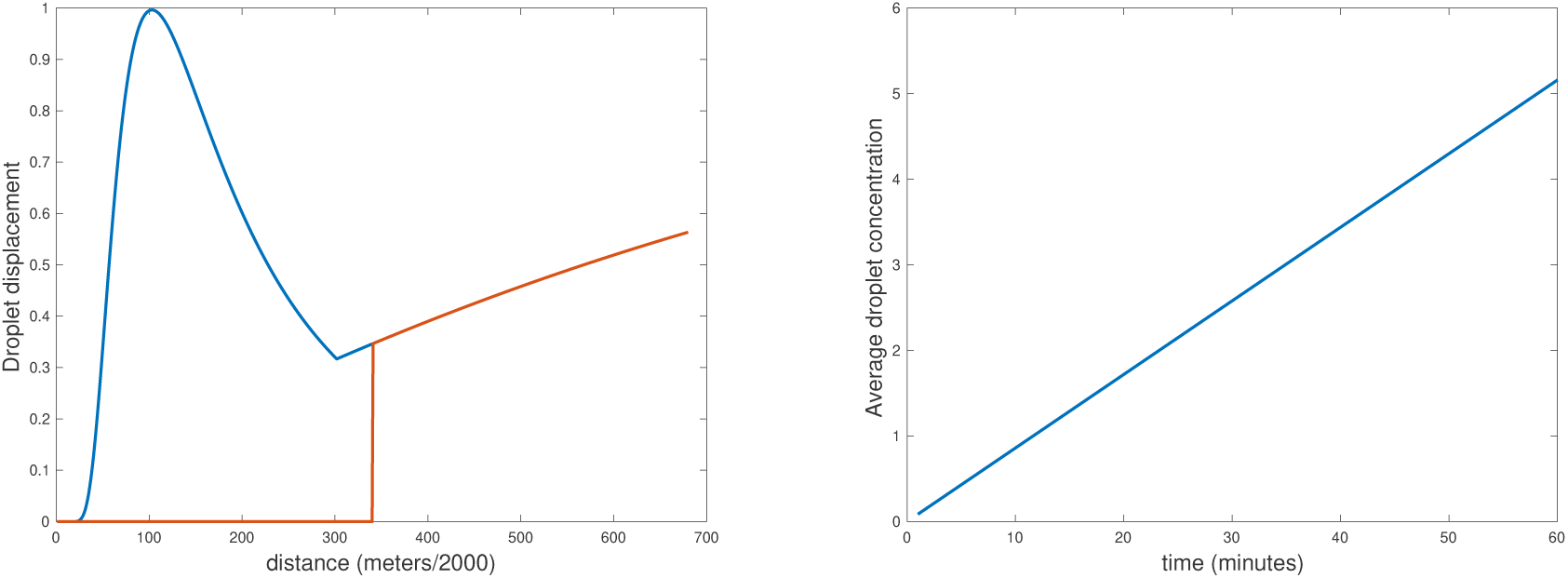
(Left) The total distribution of droplet displacement after 2.4 second of exhaling. The blue (Lagrangian) part corresponds to the first 1.2 second of the exhale event and the red (Eulerian) part the subsequent 1.2 seconds. This is a cross-cut giving the longitudinal shape of the, cylindrically symmetric, exhaled cloud. (Right) The average, in space (inside the pyramid), droplet concentration as a function of time, in minutes, normalized by the concentration at the infected person. The concentration builds up to one (the concentration at the infected person) in 12 minutes and 5.16 times that in an hour.

**Figure 4:**
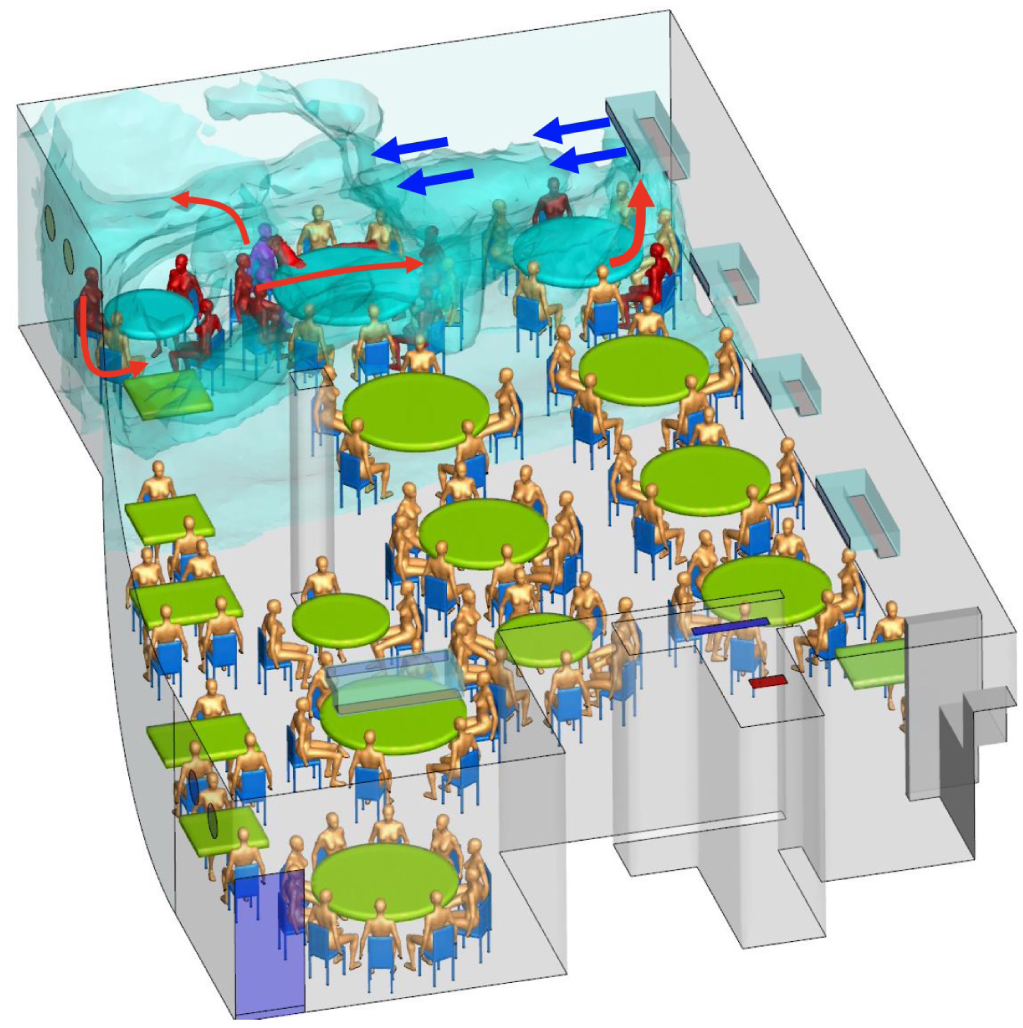
A CFD simulation of the contagion in the restaurant in Guangzhou China. The blue gas models the contamination by the droplet/aerosols. The infected people are colored red. Notice that the contaminated region roughly forms a pyramid with base on the wall opposite to the air-conditioner. The figure is take from [28].

**Figure 5:**
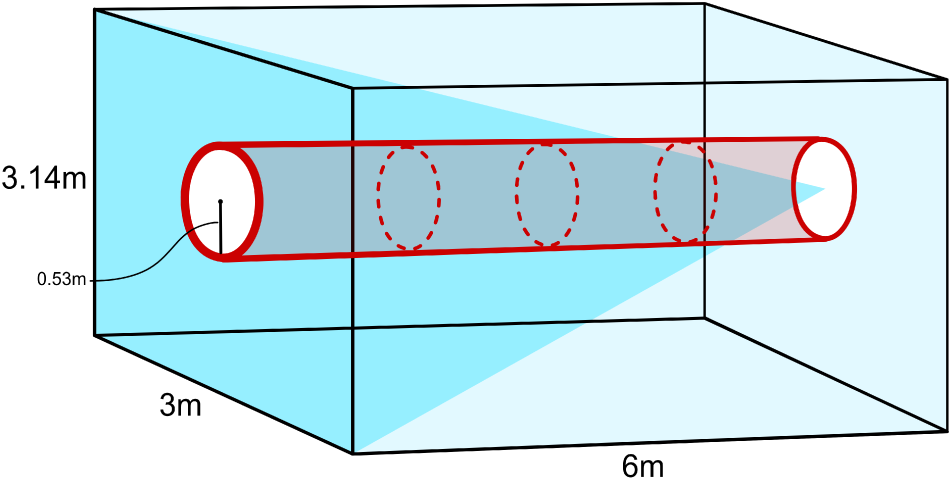
A cartoon (not to scale) of the volume (red cylinder) where the droplet/aerosol clouds propagate and the volume (blue pyramid) that the droplet/aerosol clouds get spread to by the air-conditioning.

## 3 The Concentration of Droplets, Aerosols and Virions

The conventional way of modeling the concentration C, of droplets and aerosols in a room, is by a differential equation

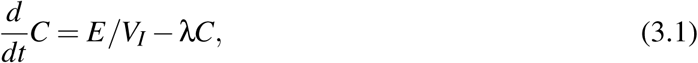

where E is the exhalation rate, in dimensions droplets and aerosols per volume per minute, and *V*_*I*_ is the volume of infectious air. In the example of the restaurant above *V*_*I*_ is the volume of the pyramid and not the volume of the whole airspace. This means that the frequently made hypothesis, that the whole available air space is well mixed, does not hold for the restaurant and may not be true in other cases. λ is the decay coefficient of the concentrations, it is composed of a sum of three decay rates λ = *k*_*v*_ + *k*_*s*_ + *k*_*d*_, where *k*_*v*_ = 1*/t*_*v*_, *k*_*s*_ = 1*/t*_*s*_, and *k*_*d*_ = 1*/t*_*d*_. *t*_*v*_ is the time it take to ventilate the room, *t*_*s*_ is the settling time of the droplet/aerosols and *t*_*d*_ the decay time of the viruses in the aerosols.

The vents in the restaurant were closed so there was no ventilation and we can take *k*_*v*_ = 0. (We elaborate on this choice in the Discussion Section.) The droplets settle over time, but they were being blown around by the air conditioner and quickly evaporate into aerosols. This process was presumably helped by modest heating. The estimates of the settling time range from 8-14 minutes, see [41] and [40], whereas the evaporation of the droplets only takes a few seconds, see [24] and [41]. Thus the settling time is just relevant for the process of turning droplets into aerosols. The settling of the aerosols is negligible, even if they settle on the tables they are being blown around by the airflow, and by restricting *C* be the concentration of aerosols, including the nuclei of the evaporated droplets, we can take *k*_*s*_ = 0. This leaves the decay time of the virions in aerosols. This time is estimated experimentally in [44], [21] and [17]. It depends on the relative humidity and is similar to that of Influenza A. Since the dependence of the latter on the relative humidity is known, we will use the decay coefficients for Influenza A [45] as a proxy. The relative humidity in Guangzhou in January is 72% but weather is cold and the relative humidity in a heated restaurant can be expected to be much less. We pick it to be 21% and use the corresponding value *k*_*d*_ = 0.0031 from [45]. Higher values will give qualitatively similar theory, but slightly different quantitatively, see Appendix D. The aerosol concentrations now lies between the two polynomials

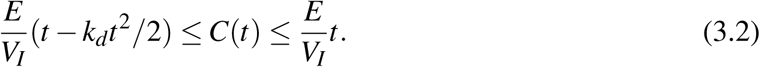

It continues to increase through the time interval and reaches its maximum *C*(60) = 4.72 at the end of the hour, see Figure 6.

**Figure 6:**
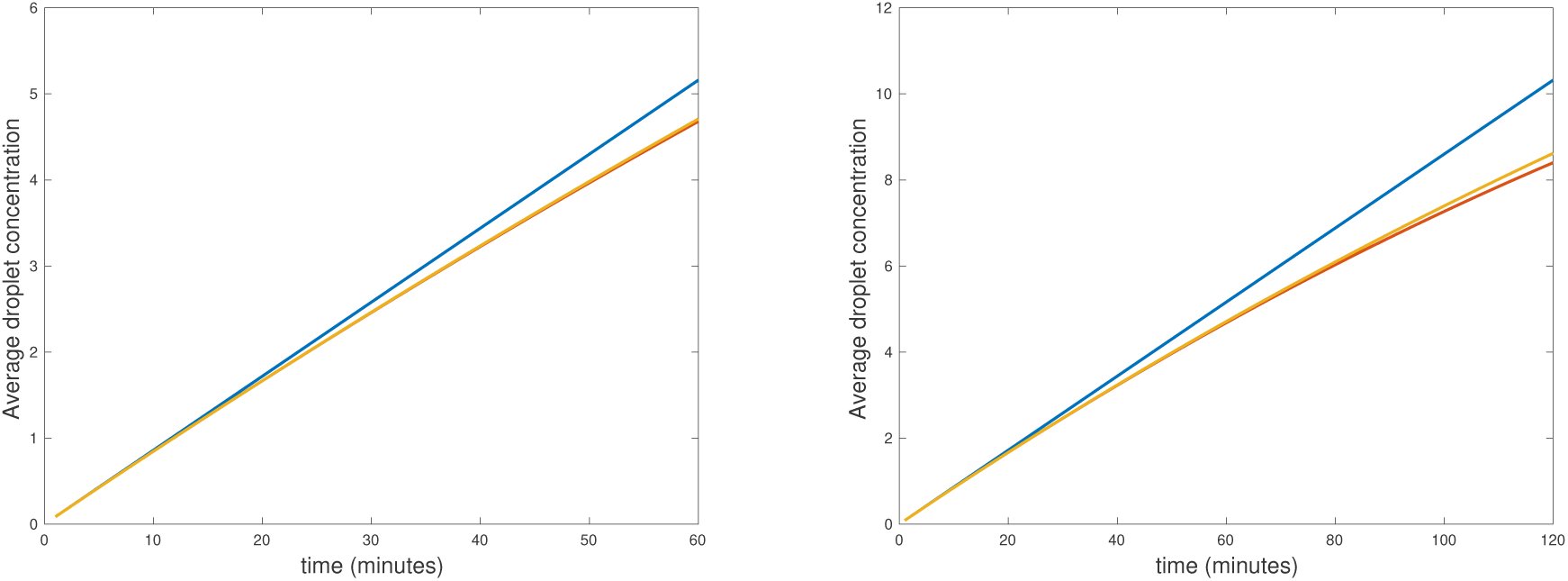
(Left) The aerosol concentration for *one hour*, top (blue) linear curve, middle (orange) solution of the ODE, bottom (red) the quadratic polynomial. The last two lines are hardly distinguishable. (Right) The aerosol concentration for *two hours*, top (blue) linear curve, middle (orange) solution of the ODE, bottom (red) the quadratic polynomial. The last two lines begin to separate on the right.

If we use the 40% relative humidity in [44] and their corresponding measured value of *k*_*d*_ = 0.01, the picture does not change much, *C*(*t*) continues to increase through the hour and reaches its maximum *C*(60) = 3.87 at the end of it, see Appendix D.

We now discuss what the concentrations of virions are represented by the aerosol concentration *C* = 1 at the infectious person, or in his or her exhaled cloud. It was shown above that the volume of the exhaled cloud after an 2.4 second exhale event is 1,320 liters. The tidal volume or the volume of the exhaled breath is 0.5 liters. Thus the concentrations of the droplets and aerosols decreases by a multiplicative factor of 0.5*/*1320 = 3.79 × 10^−4^. Now the relevant unit is *TCID*_50_ or 50% tissue-culture infectious dose. The concentration of *TCID*_50_ in the respiratory track is 10^5^ *TCID*_50_ per milliliter, see [44]. That gives 10^8^ *TCID*_50_ per liter and multiplying with the above factor, we get the concentration 3.79 × 10^4^ *TCID*_50_ per liter in the exhaled cloud. It is not yet known what the infectious dose is for people, it is higher than it is for tissue culture, but an inhale event, based on the tidal volume 0.5 liters, will involve 1.90 × 10^4^ *TCID*_50_ and if the effectiveness, of the infection, reduces the effective *TCID*_50_ by a factor of ten^1^, then there is some consensus in the literature, see [31, 11, 17, 18], that 1900 or roughly 2000, *TCID*_50_ with 50% infectious dose, is enough for infection. We can think about this as 1000 effective virions and this is what we normalize to be one, or the concentrations experienced by sitting next to the infected person, during the first couple of minutes of the outbreak event. Notice, that the concentration at the infected person may increase with time, so the normalization must be made with an average over an initial time interval of one to two minutes. In one minute a person sitting next to the infected person will have inhaled 1000 effective virions, 12 times. Consequent, people sitting next to an infectious person for a sufficiently long time (10-15 minutes) are invariably infected.

The analysis above also makes it clear that if the concentrations are higher that one, it will take a shorter time for a person in the contaminated space to become infected. In fact, the higher the concentrations, the shorter the infection time.

## 4 Discussion

In [30] it was speculated that the flow generated by the air-conditioning was blowing large droplets from table to table. This is highly unlikely since the air-conditioning flow (0.25 m/s) does not have enough energy to send the droplets on a ballistic trajectory between the tables, see [24]. In [28] the blame for the outbreak was put on the poor air-conditioning in the restaurant, caused by the closed vents and small injection of fresh air. We redid the simulation with the recommended [1] injection of fresh air 8 liters per second for each person in the infected area, to test this hypothesis. There are 21 persons at the three tables A, B and C and during the 0.83 minutes it takes the infected person to fill the cylinder above with a droplet/aerosol cloud, 8,366 liters or 8.37 *m*^3^ are injected into the infected area, in 0.83 minutes, if the air-conditioning follows recommendations. Adding this to the total volume above we get 63.81 *m*^3^, 21.27 *m*^3^ of which pass through the pyramid containing the infected air. This amounts to adding 3.36 *m*^3^ per minute, to the volume 18.48 *m*^2^ of the contaminated pyramid in Figure 5. In one hour we add 201.6 *m*^3^ of fresh air to the pyramid, for an air change per hour (ACH) equal to 10.70. This is high due to the high density of the people in the restaurant. This gives the damping coefficient *k*_*v*_ = 0.178 in Equation 3.1. The concentration now quickly increases and is 0.5, see Appendix D, in one hour. This is still high enough to cause an infection. Thus fixing the air-conditioning, significantly decreases the concentration but does not completely solve the problem. We also asked the question how much injection of fresh air would we need to make the restaurant safe? If we increase the ACH to 52.2, then *k*_*v*_ = 0.87 and this decreases the concentration to 0.1 in one hour. In other words we need more than 5 times the recommended air injection to make the contaminated pyramid have concentration less than 0.1 in one hour, see Appendix D. This amounts to a brisk breeze blowing through the restaurant.

It is clear from the above discussion that the part of the restaurant where the contagion took place was a particularly vulnerable place for transmission by aerosols. The air flow from the air-conditioner was restricting the space that needed to be ventilated, to the pyramid in Figure 5, not the whole space in the back of the restaurant. The vents were closed so there was not fresh air coming in, the gentle heating accelerated the evaporation of small drops into aerosols and the density of the patrons was very high. These factors conspired to make the ventilation coefficient *k*_*v*_ = 0 in Equation 3.1. The settling coefficient of the aerosols, that were being blown around and reinjected into the pyramid by the air-conditioner, was also *k*_*s*_ = 0. The only mitigating influence was the decay coefficient of the aerosols, that is very small *k*_*d*_ ≪ 1. The combination of these factors made the back of the restaurant in Guangzhou a very infectious place in a short time.

In [37] the transmission of the virus droplet/aerosol through an air-conditioning system is discussed with the conclusion that greater volume of outdoor air and MERV-13 or HEPA filters with the capacity of filtering our the droplet/aerosols need to be used. This is consistent with our observations. However, the air-conditioners in current use may be unable to use MERV-13 filters or handle the required volume of outdoor air. Indeed a new generation of air-conditioners that meet these requirements may be needed.

### 4.1 The problems with the conventional analysis

In this paper we have shown that the aerosol concentration in the restaurant in Guangzhou could increase as a second order polynomial in time, reaching the concentration *C* = 4.72 times what it was, *C* = 1, next to the infected person for the first couple of minutes of the outbreak event. So why has this not been discovered by previous studies ? The reasons seem to be that the previous studies are based on faulty assumptions about the conventional model, the most serious of which are the assumptions of a steady state and the assumption of a well-mixed air, see [29, 15, 20].

We discussed above how the airflow could not carry the larger droplets ballistically between the tables as speculated in [30], however the airflow can easily carry the much lighter aerosols and continue to blow them around the contaminated pyramid. There is every reason to believe that the aerosols were sucked up by the air-conditioner and distributed to the back wall were they infected the patrons at Table B, see Figure 1, since the air-conditioner did not have a filter capable of filtering out the aerosols. In [30] smear samples were collected from the air-conditioner and since those were negative it has led to the conclusion that aerosols could not have been distributed by the air-conditioner. This is clearly false, a positive result from smear samples would have confirmed the distribution of aerosols but a negative result does not show that no distribution took place. In fact, a spread of aerosols by the air-conditioner to table B is the only logical explanation of how the three people at table B got infected. The aerosols collecting on surface of the air-conditioner where probably short lived and the air-flow had plenty of time to clean them all up in the days between the outbreak and when these samples were taken. Thus the negative result from the smear samples is expected.

In [28], the concentration of tracer gas was measured but averaged over time (assuming the existence of a steady state). The time averaged concentrations at Tables B and C, were found to be close to one, but this only shows that the relative concentrations at the three tables are similar over time. It obscures the fact that the (spatially averaged) concentration at all the tables is increasing over time and the relevant reference concentration that shows this, is the concentration at Table A, see Figure 1, during the first couple of minutes of the outbreak event, see Section 3. The CFD simulation implemented in [28] show that the air in the contaminated part of the restaurant is not well mixed, but in spite of this, it assumed in [28] that the very weak airflow through elevator and fire door exhausted by the fan in the restroom, see Figure 4, is distributed throughout the restaurant. (In the experiment [28], desk fans were placed on the tables to aid the mixing.) It is very unlikely that any of this air made it to the back of the restaurant, given the strong transverse flow from all the air-conditioning units. Even, if the assumptions in [28] were correct, the quantity of air (ACH) was so small that it would amount only to (be equivalent to) a small increase in humidity, see Appendix D. The growth of the concentration would not be affected much.

The final problem that seems to have mislead some studies is the assumption of the influence of settling time for some droplets, see [20]. As we discussed above the small droplets evaporate long before they settle and their evaporation is accelerated by heating of the air by the air-conditioner.

In the most infectious cases, there may be strong airflow, without any ventilation, that reinjects the aerosols into the confined space and prevents the air in the space from being well-mixed, thus increasing the aerosol concentration further. This seems to have been the case in the restaurant in Guangzhou. The concentration is such spaces can quickly increase to very infectious levels.

## A Lagrangian Turbulence and Structure Functions

The difference between the Eulerian and the Lagrangian description of fluid flow is the frame of reference of the observer (measurements) of the flow. In the Eulerian description the observer is stationary and the fluid flows by him or her. In the Lagrangian frame of reference the observer travels with the flow and make his or her measurements in this traveling frame of reference. The mathematical description in the Eulerian frame is the conventional Navier-Stokes equation, but in the Lagrangian frame it is the Navier-Stokes equation with the material derivative 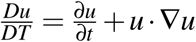 where the time variable *T* is the Lagrangian time, *u* is the fluid velocity and *t* the Eulerian time. The Lagrangian description is natural for the droplet/aerosols transmission, since the natural frame of reference is the one traveling with the droplet/aerosol, to see where it originates and where it ends up.

The Eulerian description is the traditional description of homogeneous turbulence, see [5, 25], and boundary turbulence, see [43, 9]. When particles or fluid droplets, such as the droplet/aerosols carrying the coronavirus, are entrained in the (air) flow, the Lagrangian description is more appropriate. We want to travel with (in the frame of reference of) the turbulent cloud and see how the droplet/aerosols interact with the cloud and each other. We want to determine if and where the droplet/aerosols settle on the ground or on surfaces, or if they remain airborn. In the latter case, we want to determine to what destinations they are carried by the turbulent flow of air. The strength of the turbulence in the flow is given by the dimensionless Reynolds number 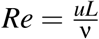, where *L* is a typical spacial dimension of the flow and ν is the kinematic viscosity. In practice, one uses the Taylor-Reynolds number 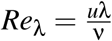, where λ is the correlation length in the flow. We will use the approximation 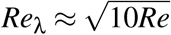.

To compute the Richardson probability density function (PDF), see Appendix B, for the droplet/aerosols that are passive scalars, we must construct the Lagrangian velocity structure functions (LVSF) [42, 2],

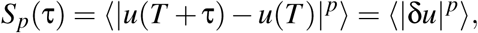

for *p* = 2, where τ is a temporal lag-variable measuring the time passed between two observations of the flow, and ⟨ · ⟩ is an ensamble average over many measurements.

The Kolmogorov-Obukhov theory determines the scaling laws in Lagrangian turbulence, for small τ,

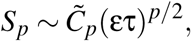

see [35, 32], where ε is the dissipation rate in the flow, and 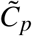 are constants, that are not universal, but depend on the configuration of the flow. In particular, for *p* = 2, we get 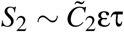, see [42]. Although the Lagrangian turbulent flow follows these scaling laws for τ sufficiently small, it quickly deviates from the predicted scaling exponents and approaches values of the scaling exponents that are more similar to their values in Eulerian turbulence [42]. In between, there is a “passover” interval where the values of the exponents dip significantly below either their initial or eventual values, see [42].

In [2], the author and his collaborators adapt the methods developed in [9], for boundary layer turbulence, to compute all the Lagrangian velocity structure functions. In [9] they had encountered a similar “passover region”. This was the buffer region separating the viscous from the inertial layer in pipe and boundary turbulence. The author and his collaborators were able to overcome this obstacle by adapting a “spectral function” introduced in [23] to compute the mean velocity in boundary and pipe flow, and generalize it to also model the buffer layer. This explains the shape of the scaling exponents observed in simulations and experiments [42]. We use these results, for *S*_2_ = ⟨|δ*u*|^2^⟩ in Lagrangian turbulence, see Figure 8, applied to the restaurant, see Figure 7.

**Figure 7:**
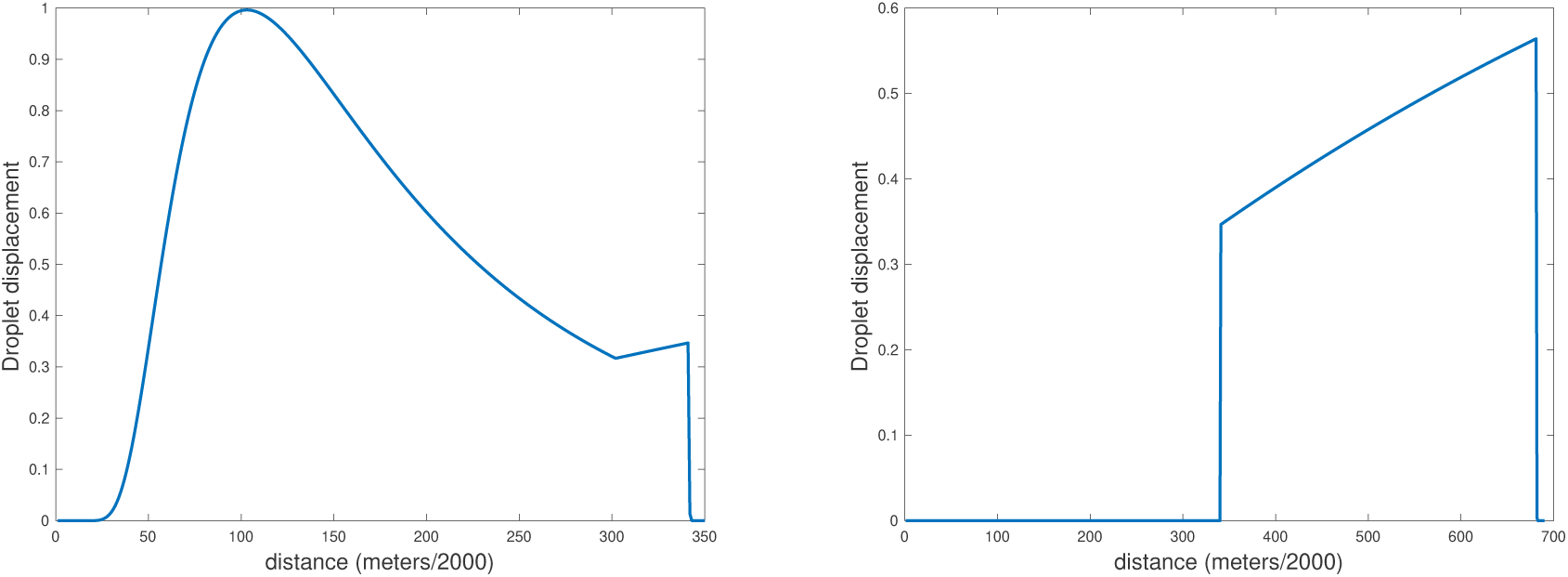
The initially exhaled (left) and subsequently propagated (right) spatial distributions of droplet displacement.

**Figure 8:**
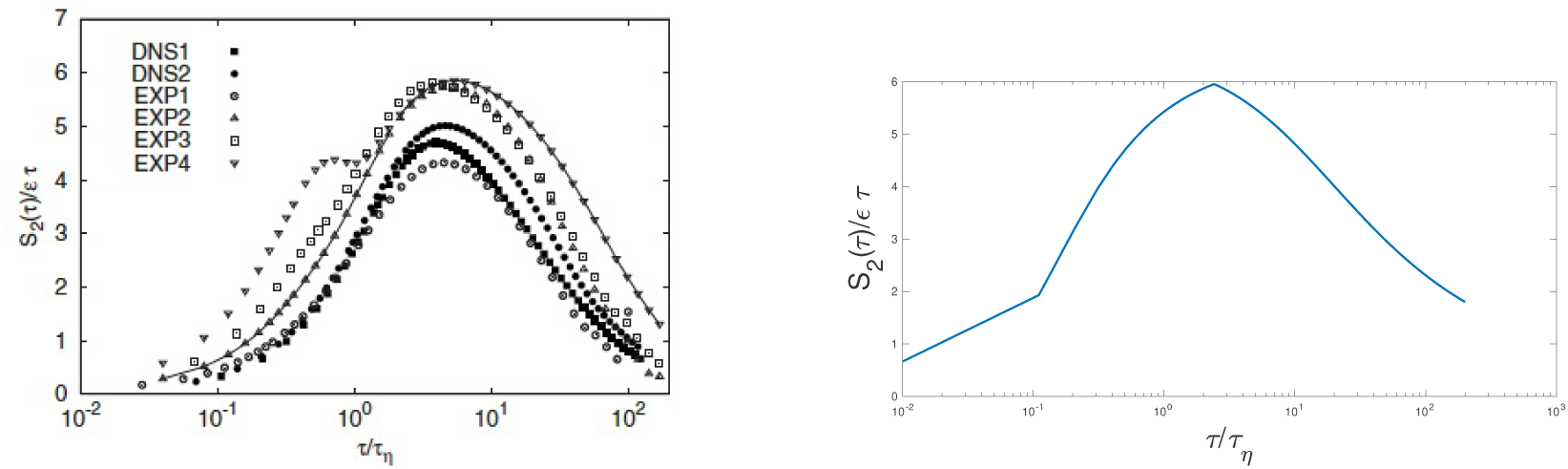
The experimental and simulated compensated second structure function *S*_2_/(ετ_η_), from [6], (left) and the theoretical model of *S*_2_/(ετ_η_) (right).

### B The Richardson Scaling

Diffusion of droplets or particles was modeled by Richardson [38] who assumed that the diffusivity is

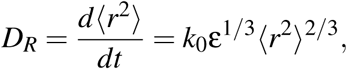

based on empirical evidence where *r*(*t*) is the particle separation at time *t*. The solution of this equation gives

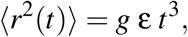

where 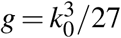 is called the Richardson constant. This is know as Richardson diffusion. Obukhov [35] gave a derivations based on the Kolmogorov-Obukhov scaling

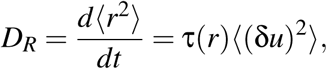

where δ*u* is the Lagrangian velocity difference, so ⟨(δ*u*)^2^⟩ is the second Lagrangian structure function and τ(*r*) is the eddy turnover time. Now according to the Kolmogorov-Obukhov Theory, ⟨(δ*u*)^2^⟩ = *C*_2_ε^2/3^*r*^2/3^ and τ(*r*) = ε^−1/3^*r*^2/3^, so

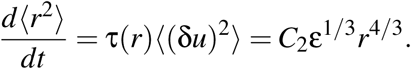

The solution is

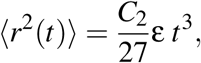

Or 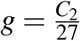. This holds for η ≪ *r*_0_ *<* ⟨*r*^2^(*t*)⟩^1*/*2^ ≪ *L*, where η is the Kolmogorov constant and *L* is the system size. *r*_*o*_ is the initial particle separation.

In addition to this region, there is a ballistic regime derived by Batchelor [4]. In this regime the particles separate linearly in *t* depending on the initial velocity. For the initial separation of the particles *r*_0_ very small, first there is a ballistic region and at a later time the particles separate exponentially. This marks the beginning of the Richardson diffusion.

We assume that the droplet/aerosols are passive scalar, or that they are simply carried along by the flow without influencing the flow itself. The probability density function, for the separation *r* of the passive scalars, satisfies the partial differential equations (PDE)

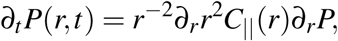

where *C*_‖_ is the longitudinal correlation function. With *C*_‖_∼ *Dr*^4*/*3^ and *P*_0_(*r, t*_0_) = δ(*t − t*_0_), the PDE has an explicit solution in the large time limit, see [27] and [3],

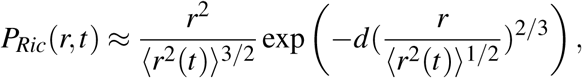

where *d* is a constant determined by *D*. This assumes that the velocity field is stochastic, incompressible, homogeneous and isotropic and δ correlated in time.

The Richardson scaling allows us to express the Richardson probability density function (PDF) in terms of the structure functions. Namely, using the above to set 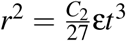, we get that 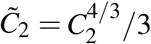, and

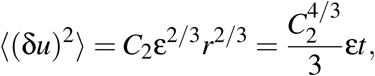

So

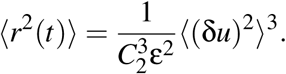

A substitution into the PDF above gives

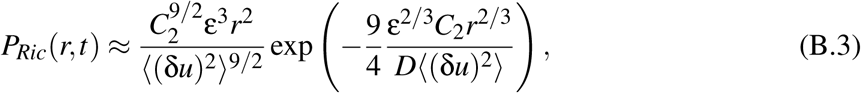

using the value 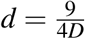 from [3]. *D* is the coefficient in the Richardson law

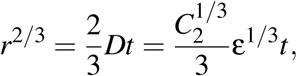

so 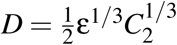.

In Figure 8, we compare the compensated second structure function *S*_2_*/*(ετ_η_), from experiments and simulations, from [6], with the theoretical model of *S*_2_*/*(ετ_η_) used in this paper.

### C Computation of the Richardson Coefficient

The second Lagrangian structure function is

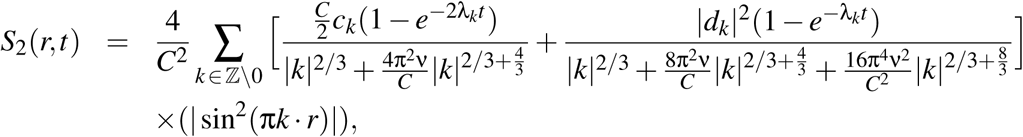

by the stochastic closure theory, see [7]. At *t* = ∞ and for *r* small we get

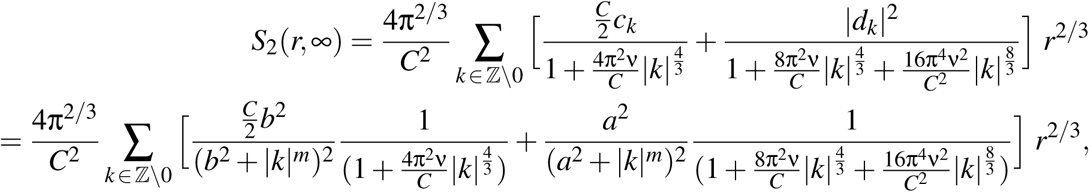

where we have used the models 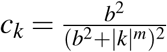 and 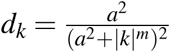 from [25], for the coefficients *c*_*k*_ and *d*_*k*_.

The Taylor-Reynolds number for the restaurant in Guangzhou is *Re*_λ_ = 705, the distance from the infected person to the wall with the air-conditioner is 3 meters, the air velocity in that direction is 0.25 m/s. The parameters *C, a, b* and *m* depend on the Reynolds number, we interpolate them from the values computed in [25], to get *C* = 5.574, *a* = 6.508, *b* = 0.076 and *m* = 1.000. The value of ε = 1.2 is obtained from [6] at *Re*_λ_ = 690, this is close to our value of 705. With this information we can compute the coefficient *C*_2_ in the structure function *S*_2_(*r*) = *C*_2_(705)*r*^2*/*3^, namely

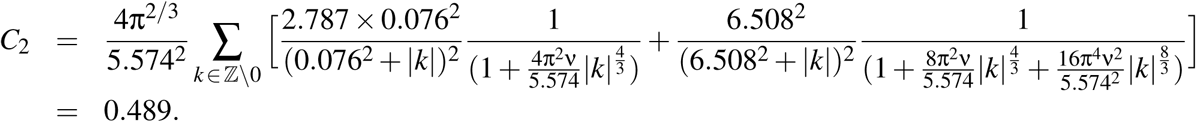

This gives the exponent in the Richardson PDF,

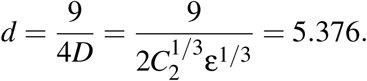

### D The Aerosol Concentration

The life-time of the SARS-CoV-2 Coronavirus in aerosols is affected by relative humidity (RH) but the exact relationship is not known yet. The dependance is presumed to be similar to that of Influenza A and we use the values for Influenza A, from [45] Table 3, as a proxy. The RH values and the corresponding values of *k*_*d*_ are given in Table 1.

**Table 1:**
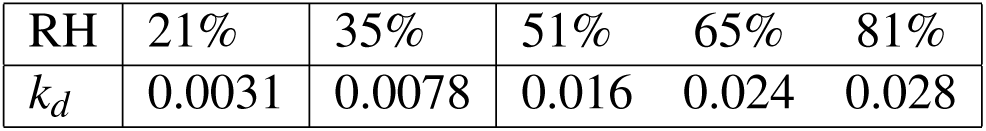
The viral decay coefficient *k*_*d*_ = 1*/time*_*d*_ for five values of relative humidity (Influenza A), from [45] Table 3.

| RH | 21% | 35% | 51% | 65% | 81% |
| --- | --- | --- | --- | --- | --- |
| $k_d$ | 0.0031 | 0.0078 | 0.016 | 0.024 | 0.028 |

In Figure 9, we show the aerosols concentrations as a function of time corresponding to the values in Table 1. The first two value of RH (left figure) give concentrations bounded below by a quadratic polynomial, the last three (right figure) give concentrations bounded below by a quintic polynomial. In all of these cases the aerosol concentration is rapidly increasing over the span of one hour.

**Figure 9:**
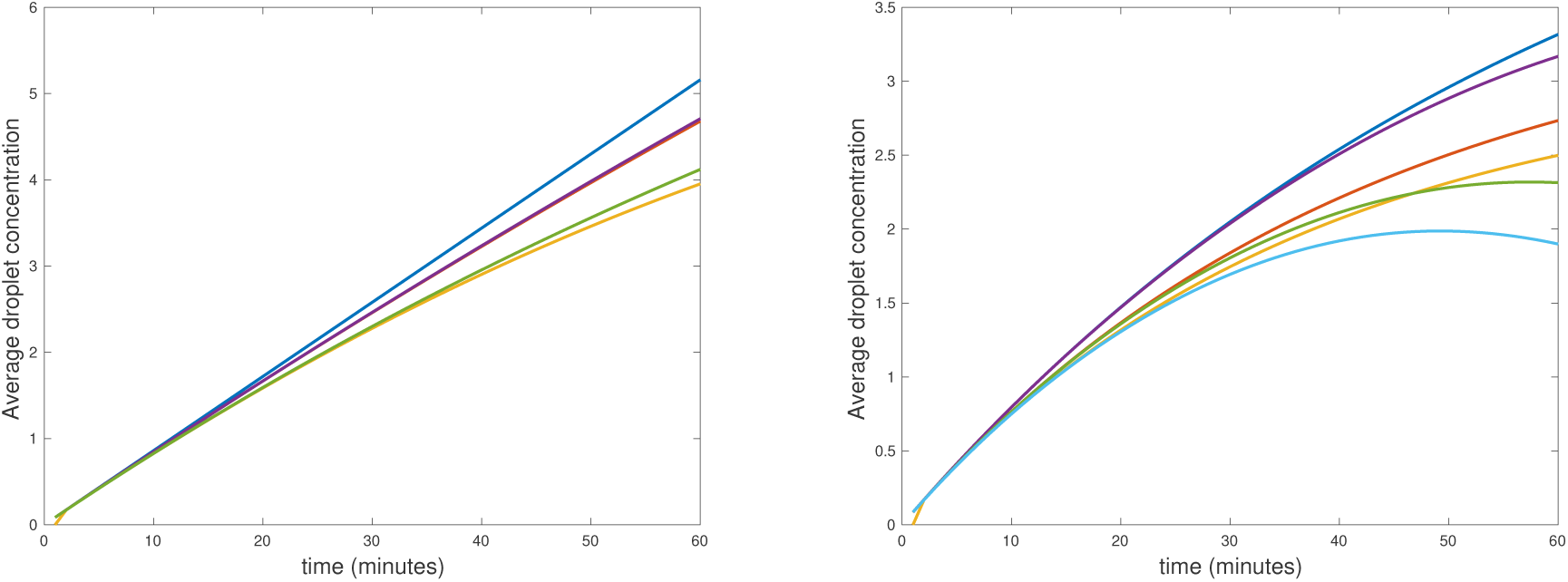
(Left) The aerosol concentration for *one hour*, top (blue) solution of the ODE, RH 21%, bounded below by a quadratic polynomial (scarlet), bottom (green) solution of the ODE, RH 35%, bounded below by a quadratic polynomial (yellow). (Right) The aerosol concentration for *one hour*, top (blue) solution of the ODE, RH 51%, bounded below by a quintic polynomial (scarlet), middle (red) solution of the ODE, RH 65%, bounded below by a quintic polynomial (green), bottom (yellow) solution of the ODE, RH 81%, bounded below by a quintic polynomial (light blue).

In Figure 10, we show the effect of ventilation. Because of the density of people in the restaurant, it is difficult to ventilate. A strong ventilation 6 ACH only reduces the concentration to 0.86 in one hour, a value that is still highly contagious. Basing the ventilation on the number of people in the contaminated part of the restaurant, 8 liters per second per person, does a little better but still leaves the concentration at almost 0.5 after one hour. This was the improvement in ventilation that we discussed in the Discussion Section above. Increasing the ventilation to 52 ACH finally brings the concentration down below the acceptable level 0.1, see the left hand side of Figure 10. On the right hand side of Figure 10 we show the aerosol concentration after one hour as a function of increasing ventilation. This figure makes it clear the restaurant is difficult to ventilate and requires 60 ACH before the contamination is mostly eliminated.

**Figure 10:**
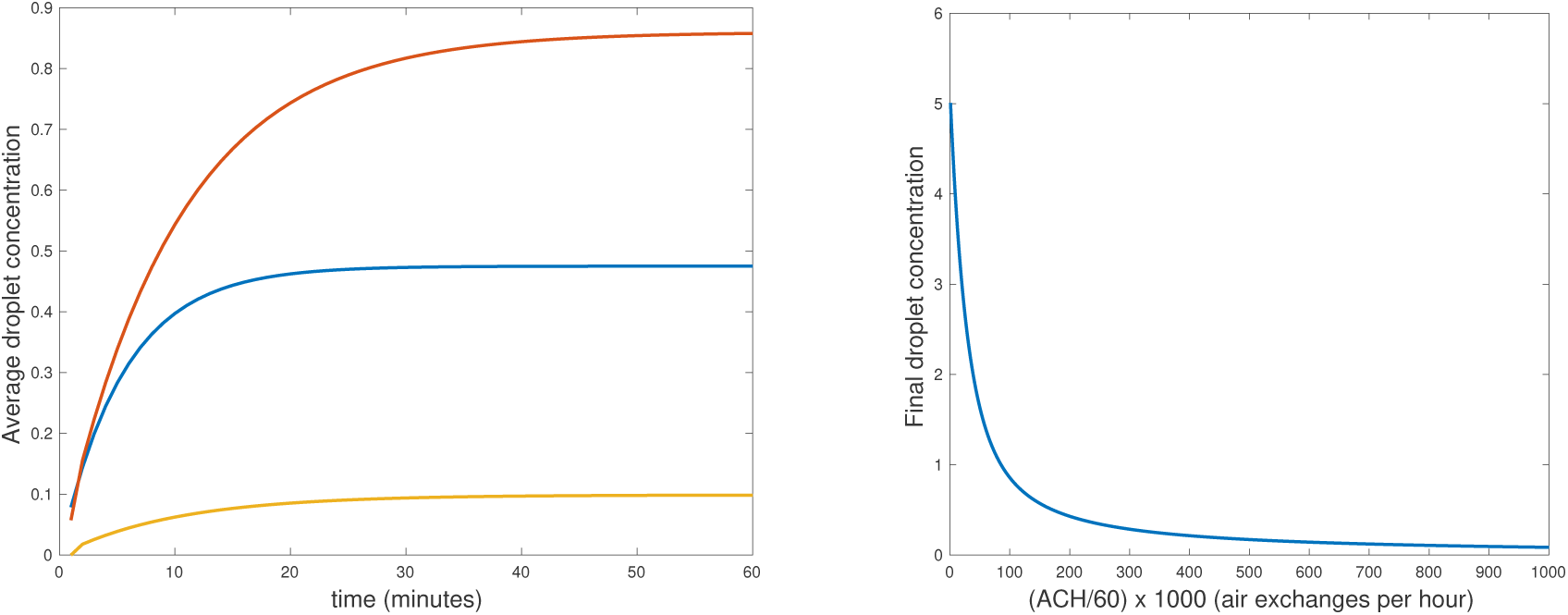
(Left) The aerosol concentration, for *one hour* with ventilation, solutions of the ODE, top (red) 6 ACH, middle (blue) 10 ACH (8 l/s per person), bottom (yellow) the ventilation that works 52 ACH. (Right) The aerosol concentration after *one hour* as a function of ventilation. The leftmost point corresponds to 60 ACH.

In Figure 11, we show the aerosol concentration as function of ventilation and time. It is clear that with little or no ventilation the concentration quickly builds up to very contagious levels but is brought sharply down by increasing the ventilation. However, to bring it to safe levels requires a very strong ventilation.

**Figure 11:**
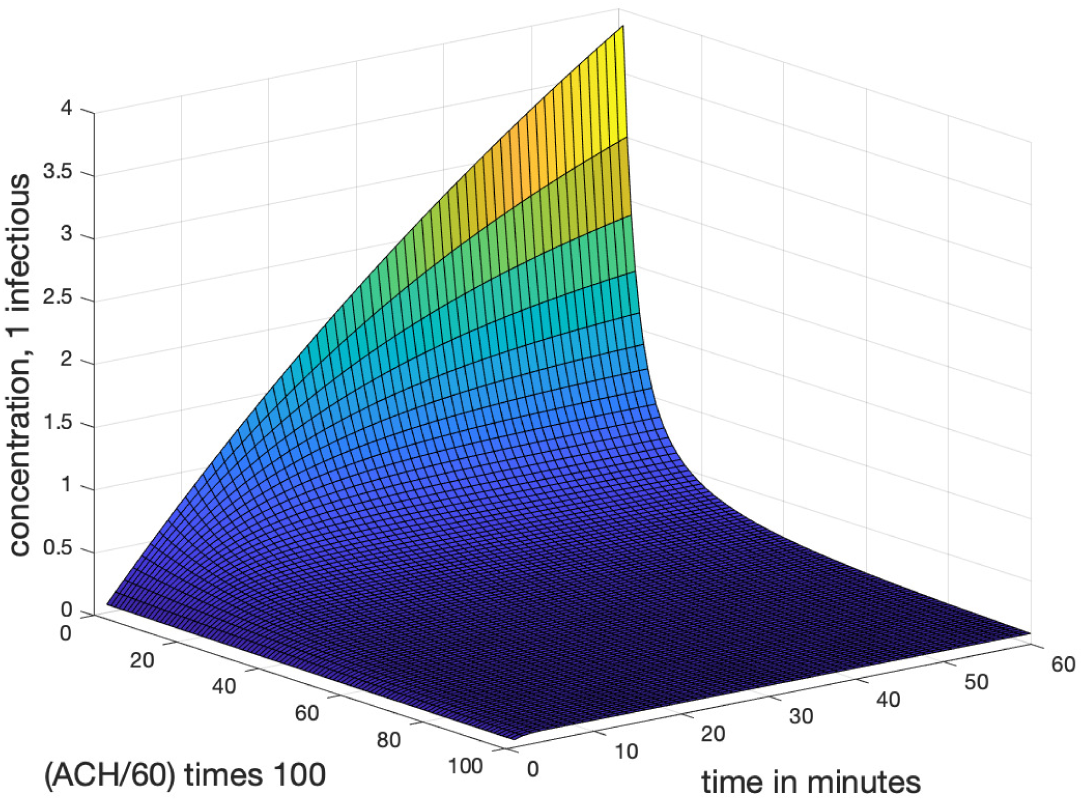
(Left) The aerosol concentration as a function of time and ventilation. The peak corresponds to poor ventilation, the low edge of the plateau to a safe (52 ACH) ventilation.

We have used used the Lagrangian computation to find the parameters in the ODE 3.1. But this was only to find the volume of the contaminated pyramid, see Figure 5. Once the Lagrangian cloud is found much more can be done. In particular, with ventilation, a configuration of restaurant tables can be found that makes next neighbor infection unlikely. In principle, we can even simulate how the aerosol cloud enters the ventilation system and is spread by it, on the average.

## Data Availability

Data is included

## Acknowledgements

The author wants to thank Luiza Angheluta, The Njord Centre, Department of Physics, University of Oslo, Norway, for helping to build the theoretical model of the structure function in Appendix A, Klaus Schauser and Albert Oaten for enthusiastic support and Ken Beisser for expert advice on air-conditioning. We also thank Knut Bauer for providing us with the graphic illustration in Figure 6.

## Footnotes

1 This factor of ten is arbitrary and just set to make connection with the consensus of 1000 virions infectious dose, that still remains to be verified.

